# Rapid Quantification of IS6110 Copy Number in *M. tuberculosis* Using Dual-Target qPCR Assay

**DOI:** 10.1101/2025.04.04.25325287

**Authors:** Daisy Patel, Krishna H. Goyani, Shalin Vaniawala, Pratap N. Mukhopadhyaya

## Abstract

PCR-based methods for detecting *Mycobacterium tuberculosis* (*M. tuberculosis*) have transformed tuberculosis (TB) diagnostics by allowing for the quick and sensitive identification of the pathogen’s DNA. The IS6110 insertion sequence is a popular molecular target due to its high specificity to the *M. tuberculosis* complex. However, the variability in IS6110 copy numbers among different strains raises questions about its reliability as a sole diagnostic marker. In this research, we devised a rapid qPCR-based technique to measure IS6110 copy numbers in clinical *M. tuberculosis* samples, comparing them with genome copy numbers estimated using a commercial MPB64-targeted kit. A custom standard curve was created using a linear DNA fragment representing the IS6110 sequence from the H37Rv strain. Analysis of 16 positive samples from 320 screened cases showed significant variability in IS6110 content. While most strains had 15–25 copies per genome, three samples had fewer than one copy per genome, likely due to mixed-strain populations dominated by IS6110-deficient bacilli. The method proved the feasibility of accurate IS6110 quantification and highlighted the importance of regional surveillance of IS6110 prevalence to prevent false negatives in PCR diagnostics. Incorporating multiple genetic targets and using dual fluorophores (e.g., FAM and VIC) in a single-tube multiplex qPCR can improve diagnostic accuracy and workflow efficiency. The findings advocate for a more nuanced, region-specific use of IS6110 in TB molecular diagnostics and emphasize the potential for automation through simple computational tools to enhance clinical decision-making.

## Introduction

PCR has become essential for the rapid and precise diagnosis of *M. tuberculosis* infections. These techniques deliver results much faster, typically within 24-48 hours, compared to culture methods that may require several weeks (Van Vollenhoven *et al*., 1996). This quick turnaround is vital for prompt diagnosis and initiation of treatment. PCR methods also exhibit high sensitivity and specificity for identifying *M. tuberculosis* DNA, even in cases with low bacterial counts, such as non-pulmonary tuberculosis (Van Vollenhoven *et al*., 1996).

Research has indicated PCR sensitivity rates as high as 98% for detecting tuberculosis patients (Beige *et al*., 1995). Notably, PCR can identify *M. tuberculosis* DNA in individuals who may not have active tuberculosis, suggesting early infection stages or asymptomatic carriers (Beige *et al*., 1995). This ability could be crucial for identifying patients at risk of developing an active disease. Although they may not entirely replace culture methods, PCR assays are gaining importance in clinical settings, particularly in resource-constrained areas, where rapid and accurate diagnosis is essential (Deggim-Messmer *et al*., 2016).

IS6110 is a widely used target for the PCR-based diagnosis of *M. tuberculosis* because of its specificity and sensitivity. This insertion sequence is repeated multiple times in the *M. tuberculosis* chromosome and is specific to the *M. tuberculosis* complex (Eisenach *et al*., 1991). PCR assays targeting IS6110 have demonstrated high sensitivity and specificity for detecting *M. tuberculosis* in various clinical samples, including sputum and cerebrospinal fluid (Deshpande *et al*., 2007; Eisenach *et al*., 1991).

However, some studies have raised concerns regarding the reliability of IS6110 as the sole target for *M. tuberculosis* detection. Variations in the IS6110 sequences among different strains of *M. tuberculosis* and the possibility of false-positive or false-negative results have been reported (Sankar *et al*., 2011). Additionally, some *M. tuberculosis* strains may lack IS6110 copies, potentially leading to false negative results (Sankar *et al*., 2011).

Despite these limitations, IS6110-based PCR remains a valuable tool for rapid *M. tuberculosis* diagnosis, often outperforming conventional methods, such as microscopy and culture, in terms of speed and sensitivity (Deshpande *et al*., 2007). However, the copy number of IS6110 varies widely among different *M. tuberculosis* lineages, with some strains carrying a single copy or lacking it. This variability poses a significant challenge in molecular diagnostics, as PCR assays that rely solely on IS6110 may fail to detect IS6110-deficient strains, leading to false negative results. Such strains have been reported in certain geographical regions, underscoring the need for a comprehensive assessment of the IS6110 copy number distribution in circulating *M. tuberculosis* populations (Howard *et al*., 1998).

In this study, we present a method for the rapid estimation of the IS6110 copy number in DNA extracted from *M. tuberculosis*-infected human clinical samples. This approach provides crucial insights into the potential underestimation of this target within *M. tuberculosis* populations, which may compromise the detection sensitivity when IS6110 is used as the sole target in PCR-based *M. tuberculosis* diagnostics.

## Material and methods

### Clinical Samples

In compliance with established biosafety and ethical guidelines, clinical samples were obtained from individuals suspected of having *M. tuberculosis* infection. Sputum samples from patients suspected of having tuberculosis were collected at the Microcare Laboratory and Tuberculosis Research Center, and subsequently processed for DNA extraction. The collection process followed institutional ethical approval, and informed consent was obtained from all the participants. To preserve sample integrity, samples were transported under controlled conditions and stored at −20°C until further analysis. The extracted DNA was then used to estimate the *M. tuberculosis* genome and IS6110 copy number, aiding the assessment of its variability in clinical *M. tuberculosis* strains.

### Confirmation of M. tuberculosis DNA and Estimation of Genome Copy Number

DNA was extracted from all clinical samples using the QIAamp DNA Mini Kit (Qiagen, Hilden, Germany), following the manufacturer’s protocol. The extracted DNA was then subjected to *M. tuberculosis* qPCR using a Commercial *M. tuberculosis* Genome Copy Number Estimation Kit (Helini Biomolecules, Chennai, India) according to the manufacturer’s instructions. Samples that tested positive for *M. tuberculosis* and generated a copy number using the kit were separated and used for further analyses.

### Obtaining Linear DNA Molecule Representing the IS6110 Segment of the M. tuberculosis Genome and Its Dilution to Generate a Quantitation Standard

DNA was isolated from the *M. tuberculosis* culture, specifically the H37Rv strain (Cole *et al*., 1998), using a QIAamp DNA Mini Kit (Qiagen, Hilden, Germany), in accordance with the manufacturer’s guidelines. This isolated DNA served as a template for amplifying the IS6110 segment, following the method outlined by Eisenach et al. (1990). The resulting amplified DNA was purified using a GeneJET PCR Purification Kit (Thermo Fisher Scientific) to eliminate contaminants and primer dimers. DNA was quantified with a Qubit 4 fluorometer (Thermo Fisher Scientific) using the corresponding Qubit assay kit. To determine the specific copy number of the IS6110 segment, the estimated DNA concentration was used to calculate the number of copies per microliter, utilizing Avogadro’s number as per the formula provided by Integrated DNA Technologies (n.d.).

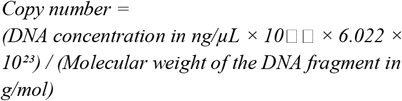

DNA was subsequently diluted to the required copy number for further application. Concentrations of 10, 10, 10, and 10 copies were used as quantification standards on a real-time PCR platform (Rotor-Gene Q, Qiagen) to create a standard curve. This curve was then used to determine the IS6110 copy number in *M. tuberculosis* DNA extracted from the clinical isolates.

### Aligning M. tuberculosis Genome Copy Number with IS6110 Copy Number

An identical volume of DNA solution was used to normalize the assays and set up a qPCR reaction to obtain the *M. tuberculosis* genome copy number (Figure 1 & Table 1), as well as the number of copies of the IS6110 repeat sequence (Figure 2 & Table 1). The latter was then divided by the former to estimate the mean IS6110 copy number per genome.

**Table 1.** Quantitative Analysis of *M. tuberculosis* Genome and IS6110 Copy Numbers in Clinical Samples.

| **Anonymised**<br>**Sample No.** | ***M. tuberculosis***<br>**genome copy**<br>**number** | **Cycle**<br>**Threshold**<br>**(Ct)** | ***M. tuberculosis***<br>**IS6110 copy**<br>**number** | **Cycle**<br>**Threshold**<br>**(Ct)** | **Computed number**<br>**of IS6110 copies per**<br>**genome of *M.***<br>***tuberculosis*** |
| --- | --- | --- | --- | --- | --- |
| DS8 | 182,369.13 | 13.97 | 6,133 | 34.02 | 0.035 |
| DS31 | 306,608.61 | 13.22 | 104,640 | 29.74 | 0.340 |
| DS20 | 417.85 | 22.82 | 795 | 37.18 | 1.90 |
| DS15 | 189.26 | 23.98 | 2,980 | 35.17 | 15.74 |

**Figure 1.**
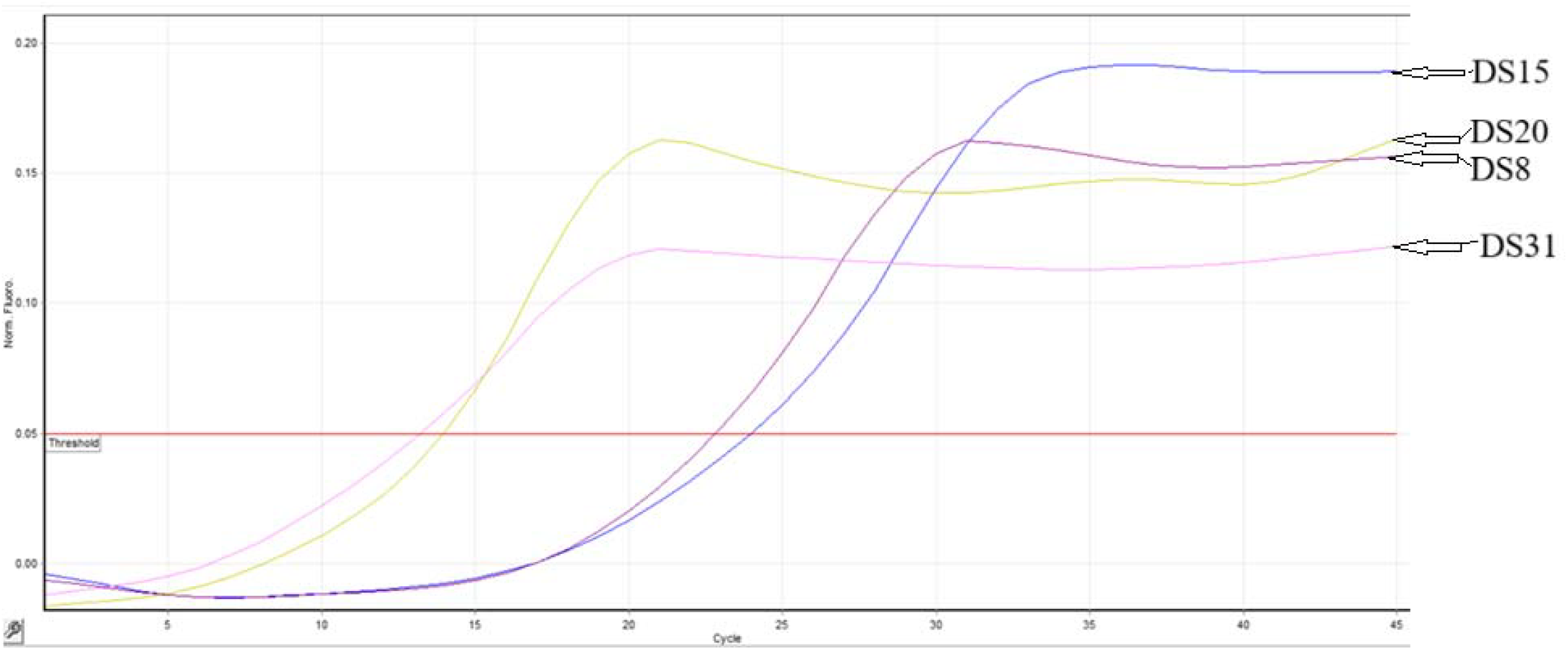
Real-time PCR (qPCR) amplification plots illustrating the cycle threshold (Ct) values for samples DS8, DS31, DS20, and DS15. These Ct values correspond to *Mycobacterium tuberculosis* genome copy numbers of 182,369.13 copies/μL (Ct: 13.97), 306,608.61 copies/μL (Ct: 13.22), 417.85 copies/μL (Ct: 22.82), and 189.26 copies/μL (Ct: 23.98), respectively. Table 1 provides a summary of the computed genome copy numbers derived from the standard curve.

**Figure 2.**
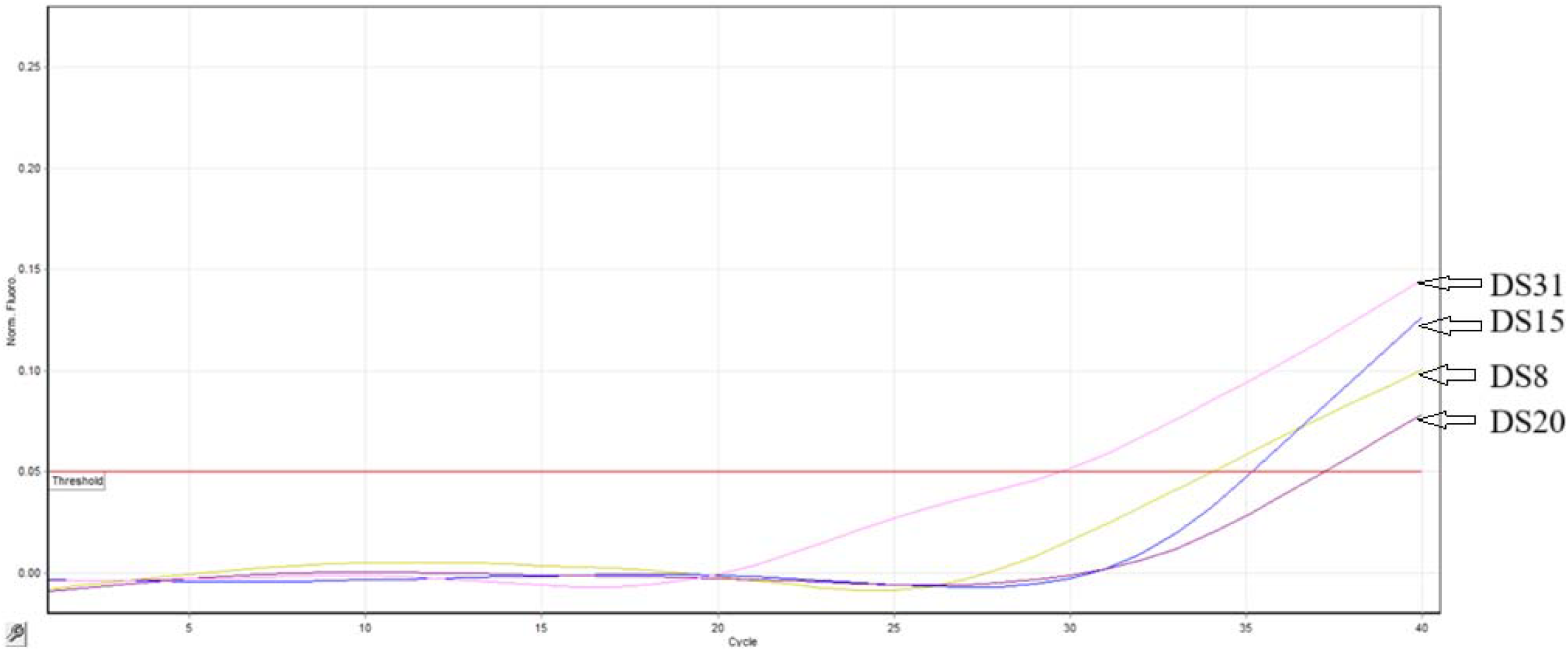
Real-time PCR (qPCR) amplification plots illustrating the cycle threshold (Ct) values for samples DS8, DS31, DS20, and DS15. These Ct values correspond to *Mycobacterium tuberculosis* IS6110 Insertion Sequence copy numbers of 6333 copies/μL (Ct: 34.02), 104640 copies/μL (Ct: 29.74), 795 copies/μL (Ct: 37.18), and 2980 copies/μL (Ct: 35.17), respectively. Table 1 provides a summary of the computed genome copy numbers derived from the standard curve.

### Ethics Statement

This study was approved by the Wobble Base Bioresearch Ethics Committee (Approval No. WBBPL/EC/Mar2025/006) was conducted in accordance with the ethical guidelines outlined in the Declaration of Helsinki. Written informed consent was obtained from all participants or their legal guardians before sample collection and analysis.

### Age Statement

The participants in this study ranged in age from 15 to 65 years, encompassing a population at a significant risk for *M. tuberculosis* infection. This range includes young adults, who represent a high-burden group for tuberculosis, as well as older adults, who may present with chronic or reactivated infections. To ensure confidentiality, age data were reported at five-year intervals.

## Results

Samples of *M. tuberculosis* were obtained from a laboratory certified by the National Tuberculosis Elimination Program (NTEP) for *M. tuberculosis* diagnostics. Of the total, 16 samples, accounting for approximately 5% of the samples screened (n=320), were found to contain *M. tuberculosis* DNA when tested using a commercial qPCR detection kit designed to identify a single-copy gene (MPB64) within the genome of the pathogen. In addition to identifying the *M. tuberculosis* genome, this kit facilitates the estimation of the *M. tuberculosis* genome copy number through a standard curve created using the kit’s provided copy number standards (Figure 1). The IS6110 copy number was estimated using an equivalent volume of extracted DNA and the copy number standard developed in this study (Figure 2). In three samples that tested positive for *M. tuberculosis*, the calculated IS6110 copies per genome were less than one. Table 1 presents the results. Here, DS15 represents one of the 13 *M. tuberculosis*-positive samples, where the IS6110 copy number ranged from 15 to 25 per genome.

## Discussion

The anticipated positivity rate for *M. tuberculosis* in this study was approximately 5%. This aligns with the standard benchmark observed in numerous NTEP-led active case-finding efforts nationwide (Burugina Nagaraja *et al*., 2021).

Molecular PCR-based methods for detecting *M. tuberculosis* have become a major breakthrough in the diagnosis of tuberculosis, offering several key benefits over traditional techniques. PCR allows for quick, sensitive, and specific identification of *M. tuberculosis*, facilitating a more rapid diagnosis and the start of treatment (Kox *et al*., 1995). Unlike conventional culture methods, which may require weeks, PCR can identify *M. tuberculosis* within 48 h (Eisenach *et al*., 1991). Generally, PCR is more sensitive than microscopy, with one study indicating 48% sensitivity for PCR compared to 9% for microscopy in cerebrospinal fluid samples (Kox *et al*., 1995). Notably, although PCR is promising, its effectiveness can differ depending on the sample type and study environment. For respiratory samples, one study reported 100% sensitivity and specificity, whereas for non-respiratory samples, the sensitivity and specificity were 85.7% and 97.3%, respectively (Malbruny *et al*., 2011). Some studies have shown lower sensitivities than culture methods, emphasizing the need for further refinement (Shawar *et al*., 1993).

Therefore, PCR-based detection of *M. tuberculosis* provides significant benefits over traditional methods in terms of speed, sensitivity, and specificity. It has the potential to enhance early diagnosis, particularly in resource-constrained settings where quick detection is vital (Salimiyan Rizi *et al*., 2020).

IS6110 is a widely used target for detecting *M. tuberculosis* using PCR because of its specificity and sensitivity. The insertion sequence IS6110 belongs to the IS3 family and is considered to be specific to the *M. tuberculosis* complex (Sankar *et al*., 2011). It has been extensively used for laboratory detection of tuberculosis and epidemiological investigations (Sankar *et al*., 2011). Several studies have demonstrated the effectiveness of IS6110-based PCR. For instance, a PCR assay targeting a 123-bp segment of IS6110 showed high sensitivity and specificity for detecting *M. tuberculosis* in sputum samples within 48 h (Eisenach *et al*., 1991). Another study using a 317-bp segment of IS6110 reported sensitivities of 55-74% and specificities of 95-98% compared to culture methods (Shawar *et al*., 1993). IS6110-based PCR has also been successful in detecting *M. tuberculosis* in paraffin-embedded skin biopsies from patients with cutaneous tuberculosis, showing higher positivity (90%) than culture methods (65%) (Margall *et al*., 1996). However, concerns have been raised regarding the specificity and sensitivity of IS6110-based PCR. False-negative results owing to the absence of IS6110 copies in some strains have been reported (Sankar *et al*., 2011).

While IS6110 remains a popular and effective target for *M. tuberculosis* detection by PCR, researchers have suggested developing multiplex PCR assays targeting multiple genomic regions to improve accuracy and reliability (Sankar *et al*., 2011). The IS6110 sequence shows variations among different *M. tuberculosis* strains, emphasizing the need for careful primer design and assay optimization (Sankar *et al*., 2011).

Various PCR-based techniques have been devised to identify and amplify IS6110 sequences, such as one-tube nested PCR (Kent *et al*., 1995) and strand displacement- and PCR-based amplification that focuses on specific IS6110 regions (Hellyer *et al*., 1996). These approaches have demonstrated high sensitivity and specificity for detecting *M. tuberculosis* complexes. Notably, (Kolk *et al*., 1994) outlined a modified target created by inserting 56 nucleotides into the IS6110 element of *M. bovis* BCG and incorporating it into *M. smegmatis*. This altered strain served as an internal control to identify issues in DNA isolation or the presence of inhibitors in clinical samples. Furthermore, it facilitates the estimation of IS6110 copy numbers in clinical samples by allowing competition between different target DNAs. Although not a commercial kit, (Plikaytis *et al*., 1993) presented a new gene amplification technique known as IS6110-ampliprinting, which evaluates the variability of the IS6110 insertion sites. This method can cluster epidemiologically related *M. tuberculosis* strains and is directly applicable to sputum specimens and *M. tuberculosis* cultures. However, it should be noted that there is no simple kit or protocol available for quantifying IS6110 in *M. tuberculosis* DNA.

In this study, we introduced a swift and effective technique for measuring the IS6110 insertion sequence in the *M. tuberculosis* genome. This method allows for the assessment of variability in IS6110 copy numbers using a polymerase chain reaction (PCR) assay that concurrently identsifies *M. tuberculosis* and determines the IS6110 copy count. By calculating the ratio of IS6110 copies to genome copies from the assay, we could precisely ascertain the IS6110 copy number per genome.

Sample DS15 showed an IS6110 copy number of about 16 per genome. This sample was part of a group of 16 *M. tuberculosis*-positive samples identified in this study, where the IS6110 copy number per genome varied from 15 to 25. This estimate was obtained by determining the ratio of *M. tuberculosis* genome copies to the corresponding IS6110 insertion sequence copies. Notably, when the same calculation was applied to samples DS8 and DS31, the average IS6110 copy numbers per genome were approximately 0.035 and 0.34, respectively. This is likely due to the presence of mixed strains in these clinical samples, where a very small population contained IS6110 elements. These IS6110-positive bacilli were spread across a large population of IS6110-deficient bacilli, leading to a significantly low average number of IS6110 copies per genome. In contrast, sample DS20 showed a slightly higher IS6110 insertion sequence count per genome, with an average calculated value of about 1.9.

In India, *M. tuberculosis* strains with only 1-2 copies of IS6110 are commonly found. These strains present difficulties for molecular epidemiological studies that rely on IS6110-based fingerprinting because of their limited ability to differentiate between strains (Bauer et al., 1999). To enhance strain differentiation, additional genotyping techniques such as spoligotyping are frequently used alongside IS6110 RFLP for strains with low copy numbers (Bauer et al., 1999). This combined approach improves the capacity to distinguish between strains and offers more precise insights into the epidemiology of tuberculosis in areas where low-copy-number strains are prevalent.

In this study, the same fluorophores have been employed to detect both targets: MPB64 for genome estimation and IS6110 for insertion sequence quantification. However, the use of different fluorophores, such as FAM and VIC, can simplify the detection process into a single-tube reaction. In such a configuration, the *M. tuberculosis* genome copy number can be identified in one fluorescence channel, while the IS6110 copy number can be detected in another, improving the assay efficiency and minimizing the risk of cross-reactivity. A simple Python script can also be created to automate the calculation of IS6110 copies within the *M. tuberculosis* genome.

This protocol can facilitate the determination of IS6110 in *M. tuberculosis* strains from specific geographical areas, assisting in evaluating the appropriateness of IS6110 as a molecular target for PCR-based detection methods. Notably, the IS6110 copy number varied among *M. tuberculosis* strains, with some strains having few or no copies of this insertion sequence. Therefore, understanding the regional distribution of IS6110 is essential for optimizing the diagnostic strategies. Additionally, incorporating multiple targets, such as combining IS6110 with MPB64, has been shown to enhance the sensitivity and specificity of PCR assays for the detection of *M. tuberculosis*. This multiplex approach ensures robust detection across various *M. tuberculosis* strains, including those with low or absent IS6110 copies, thereby improving the diagnostic accuracy in different epidemiological contexts.

## Conclusion

This study presents a rapid PCR-based technique for measuring the IS6110 insertion sequence within the *M. tuberculosis* genome, allowing for the evaluation of copy number variation. Same fluorophore was used to measure MPB64 and IS6110 in a two-tube setup. However, using different fluorophores, such as FAM and VIC, the method can be modified to detect *M. tuberculosis* genome copies and IS6110 copies simultaneously in a single-tube reaction, thus improving diagnostic efficiency. A straightforward Python script can further automate the calculation of IS6110 copies per genome, thereby offering valuable insights into the regional distribution of this insertion sequence. These data are essential for assessing the utility of IS6110 as a molecular target in PCR-based detection methods, particularly given the copy number variation among different *M. tuberculosis* strains.

## Data Availability

https://drive.google.com/drive/folders/1xnNxor8pZkifEOL27Ui43OUOTJIldGhO?usp=drive_link.

## Data Access Statement

Research data supporting this publication are available from the Wobble Base repository at located at https://drive.google.com/drive/folders/1xnNxor8pZkifEOL27Ui43OUOTJIldGhO?usp=drive_link.

## Author Contributions

DP maintained the *M. tuberculosis* DNA bank and performed the *M. tuberculosis*-specific assays using the thermal cycler. KHG developed the *M. tuberculosis* IS6110 quantitation standards for estimating IS6110 insertion sequence copy numbers. SV contributed to the research design. PNM conceived the original idea and supervised the project

## Acknowledgement

The authors express their gratitude to Microcare Laboratory and Tuberculosis Research Center, Surat, for providing the clinical samples essential for this study.

